# Efficacy of colchicine with or without corticosteroids in hospitalized COVID-19 patients: A systematic review and meta-analysis

**DOI:** 10.1101/2025.04.04.25325283

**Authors:** Marisa D. Arthur, Jason Wilson, Enze Cai, JongWon See, Brent M. Peterson, Thushara Galbadage

**Author notes:** Now in the Department of Applied Health Sciences, Texas Christian University, Fort Worth, TX, US.

## Abstract

**Background:** Colchicine, an anti-inflammatory drug, was used early in the COVID-19 pandemic for its ability to modulate the inflammatory response. However, its clinical efficacy in hospitalized patients, particularly when used alongside corticosteroids, remains unclear. This systematic review and meta-analysis assessed colchicine’s efficacy in hospitalized COVID-19 patients.

**Methods:** We searched PubMed, Web of Science, and EMBASE databases for randomized controlled trials (RCTs) evaluating colchicine in hospitalized adults with moderate-to-severe COVID-19. Risk of bias was assessed using the Cochrane tool. A random-effects model was used for meta-analysis, and heterogeneity was assessed using the I² statistic.

**Results:** Thirteen RCTs involving 16,529 participants were included in the meta-analysis. Colchicine did not significantly reduce overall mortality (Risk Difference (RD) = −0.02, 95% Confidence Interval (CI): [-0.04, 0.00]), ICU admissions (RD = −0.03, 95% CI: [-0.06, 0.01]), mechanical ventilation rates (RD = −0.01, 95% CI: [-0.02, 0.00]), hospitalization duration (Mean Difference (MD) = −0.58 days, 95% CI: [-1.80, 0.64]), or ICU length of stay (MD = −1.41 days, 95% CI: [-3.35, 0.53]), although in every comparison colchicine treated groups had a greater positive effect. Subgroup analysis showed a significant reduction in mortality among patients who received colchicine in the absence of corticosteroids (RD = −0.07, 95% CI: [−0.11, −0.03]), whereas no significant effect was observed in patients treated with colchicine and corticosteroids.

**Conclusions:** While colchicine did not improve outcomes in the overall population, it showed benefits in COVID-19 patients not receiving corticosteroids. These findings suggest a potential role for colchicine in targeted clinical settings.

**GRAPHICAL ABSTRACT:** 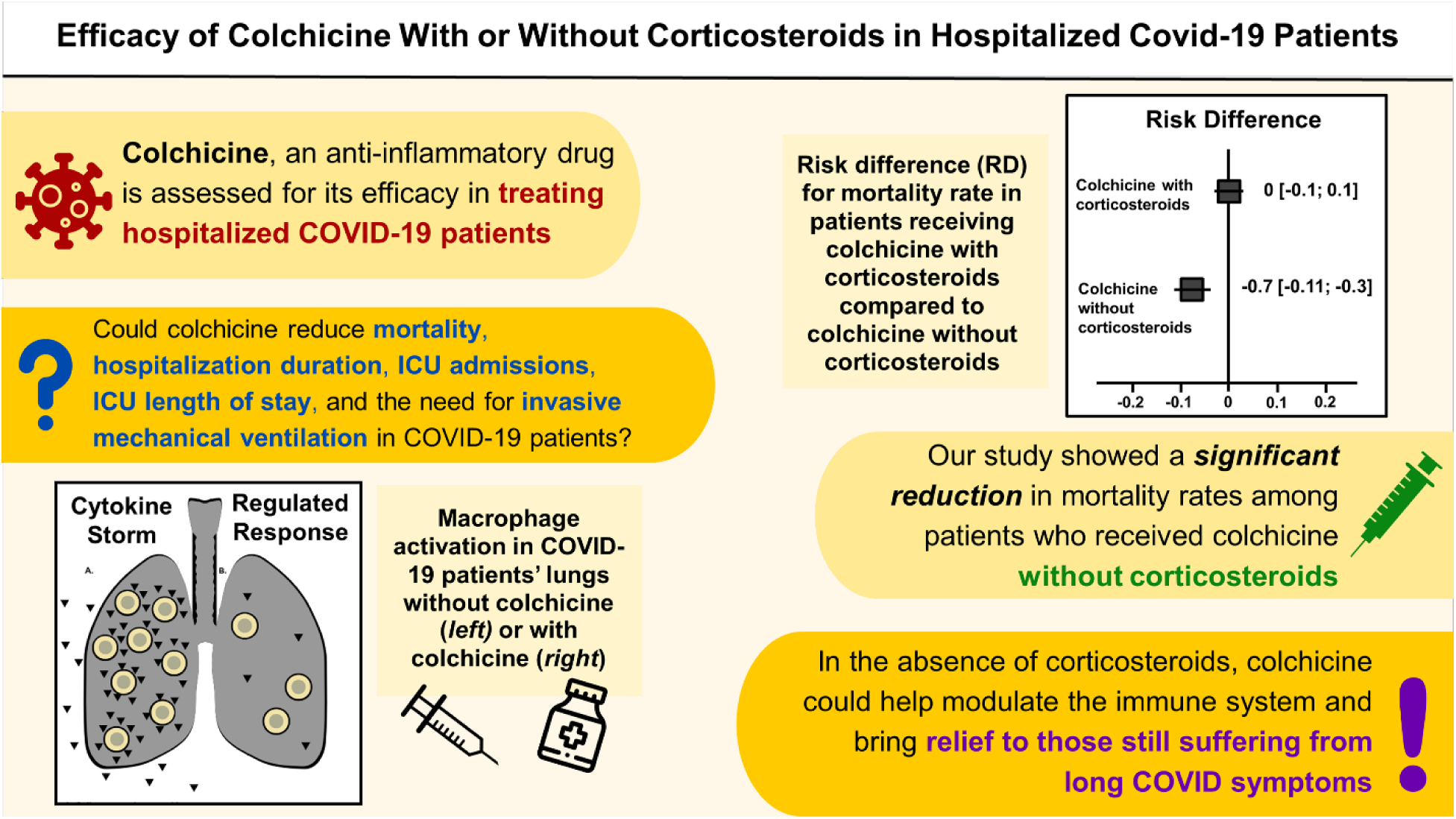

## INTRODUCTION

The COVID-19 pandemic has caused widespread morbidity and mortality, with over 7 million hospitalizations and 1.1 million related deaths reported in the United States alone as of 2024 (CDC, 2024). While vaccines may influence the severity reduction of infection, effective treatment strategies remain critical, especially for hospitalized patients with moderate-to-severe disease (NobelPrize.org, 2023; WHO, 2024). The excessive immune response (cytokine storm) triggered by SARS-CoV-2 infection plays a major role in severe cases, leading to acute respiratory distress syndrome (ARDS), multi-organ dysfunction, and increased mortality (Montazersaheb et al., 2022) (Figure 1a). Anti-inflammatory therapies such as corticosteroids have been widely adopted as part of the standard of care, yet alternative or adjunct therapies are needed to further optimize treatment outcomes (Wagner et al., 2021).

**Figure 1.**
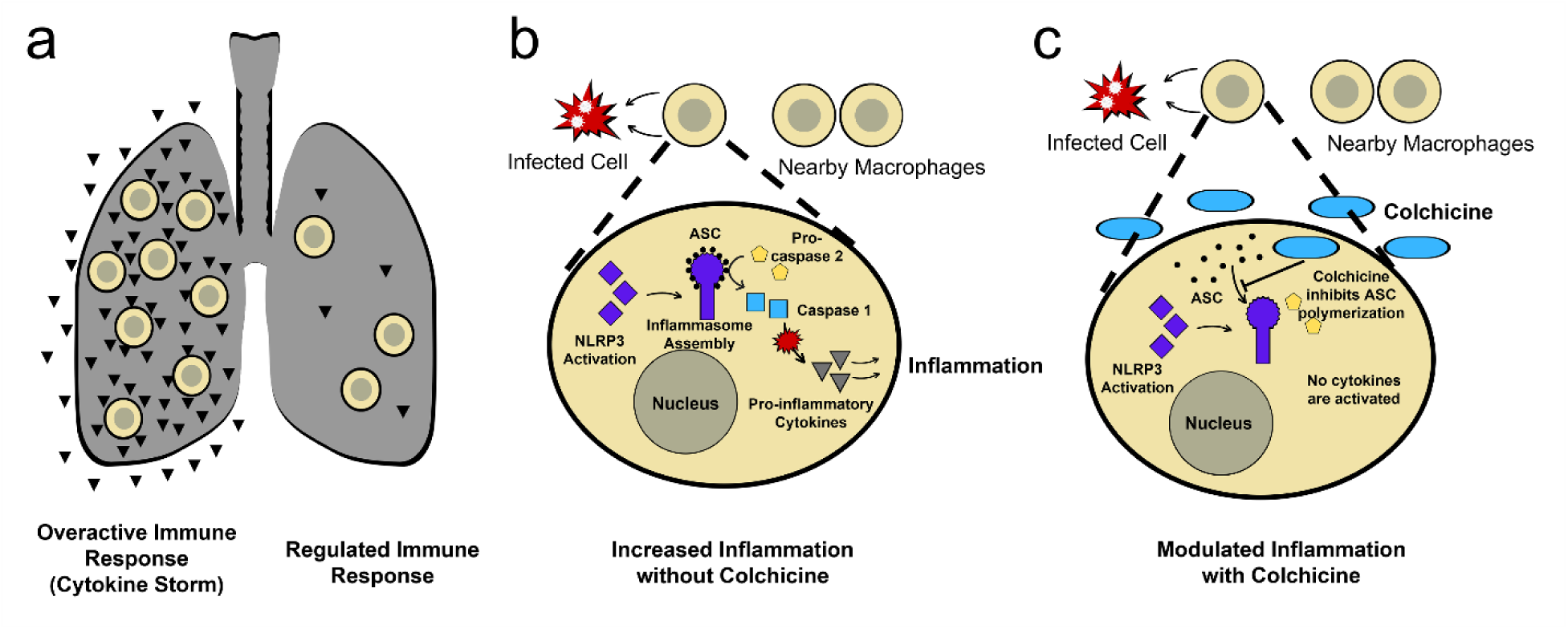
Mechanism of action of colchicine in modulating the immune response. (a) Overview of lung inflammation in response to COVID-19. *Left lung:* Uncontrolled cytokine production by macrophages and other immune cells leads to excessive inflammation, characteristic of a cytokine storm. *Right lung:* A regulated immune response with balanced cytokine production and mechanisms in place to control inflammation. (b) Macrophage activation in response to SARS-CoV-2 infection in the absence of colchicine. The NLRP3 inflammasome assembles, recruiting ASC adapter proteins and activating caspase-1, which promotes the release of pro-inflammatory cytokines such as IL-1β and IL-6, leading to an amplified inflammatory response. (c) Macrophage activation in the presence of colchicine. Colchicine inhibits the polymerization of ASC adapter proteins, preventing caspase-1 activation and subsequent cytokine release, thereby reducing inflammation.

Colchicine, a widely used anti-inflammatory drug for gout, was investigated as a potential treatment for COVID-19 due to its ability to inhibit the NLRP3 inflammasome, suppress neutrophil migration, and reduce pro-inflammatory cytokine production (Bonaventura et al., 2022). The activation of the NLRP3 inflammasome plays a key role in cytokine storm and contributes to systemic inflammation and lung injury. By interfering with microtubule polymerization, colchicine not only disrupts the inflammatory cascade but also stabilizes endothelial cells, potentially reducing the risk of thromboembolic complications, which are common in COVID-19 (Marques-da-Silva et al., 2011) (Figure 1b and c). These mechanisms suggest that colchicine could be beneficial in mitigating the hyperinflammatory response associated with severe disease. However, its clinical efficacy in hospitalized COVID-19 patients remains uncertain, particularly when administered alongside corticosteroids.

Several systematic reviews and meta-analyses have previously evaluated colchicine’s efficacy in COVID-19, but their findings have been inconsistent. While some studies have reported reductions in mortality and hospitalization duration, others have found no significant benefit (Supplemental Table 1). In addition, previous systematic reviews focusing on colchicine were conducted earlier in the pandemic and did not include more recent studies (Supplemental Table 2). Another limitation of previous reviews is the lack of stratification based on corticosteroid use. Given that corticosteroids are a standard component of COVID-19 treatment, their potent anti-inflammatory effects may obscure or modulate colchicine’s efficacy, making it difficult to determine whether colchicine provides independent therapeutic benefits. By addressing these gaps, this systematic review aims to provide a clearer understanding of colchicine’s role in hospitalized COVID-19 patients. This systematic review and meta-analysis evaluate the efficacy of colchicine in reducing mortality, hospitalization duration, ICU admissions, ICU length of stay, and the need for invasive mechanical ventilation in hospitalized COVID-19 patients. A key focus of this study was to assess whether colchicine’s effectiveness varies based on corticosteroid co-administration and treatment duration, helping to refine its potential role in clinical practice.

## METHODS

This systematic review and meta-analysis adhered to the Preferred Reporting Items for Systematic Reviews and Meta-Analyses (PRISMA) 2020 guidelines (Page et al., 2021) and was registered in PROSPERO (Registration ID: CRD42024510295) (https://www.crd.york.ac.uk/PROSPERO/view/CRD42024510295).

### Eligibility Criteria

Studies were included if they met the following criteria: participants were hospitalized adults (≥ 18 years) diagnosed with moderate-to-severe COVID-19 based on the definitions provided in each study. The intervention was colchicine therapy, either alone or in combination with standard treatments, while the comparator group received standard of care (SoC) with or without a placebo. Only randomized controlled trials (RCTs) were included, and studies had to report at least one of the primary outcomes, which included mortality rate, ICU admission rates, need for invasive mechanical ventilation, hospitalization duration, and ICU length of stay. Secondary outcomes included BMI, sex distribution, colchicine dosage and duration, adverse events, and corticosteroid co-administration. Studies were excluded if they focused on pediatric patients, non-hospitalized individuals, patients with mild COVID-19, or observational study designs. Studies published in languages other than English and those not reporting at least one primary outcome were also excluded.

### Sources and Search Strategy

A comprehensive search was conducted across PubMed, EMBASE, and Web of Science from February 2024 to April 2024. The search strategy utilized keywords including colchicine, COVID, SARS-CoV-2, hospitalized, and clinical trial. A detailed list of the search terms and results for each database is provided in Supplemental Table 3. Additional studies were identified by screening the reference lists of previous systematic reviews on colchicine in COVID-19 treatment. All retrieved records were imported into Excel for duplicate removal.

### Selection Process

The study selection followed a two-stage screening process conducted by two independent reviewers. In the first stage, titles and abstracts were screened for eligibility, ensuring that studies included hospitalized COVID-19 patients and reported at least one primary outcome. In the second stage, full-text articles from the remaining studies were assessed to confirm eligibility. Any discrepancies in study selection were resolved through discussion with a third reviewer, and reasons for exclusion at the full-text stage were documented.

### Data Collection and Extraction

Data extraction was performed independently by two reviewers using a standardized data extraction form created in Google Sheets. Extracted data included study characteristics (authors, year, country, registration ID), participant demographics (age, sex, BMI, comorbidities if available), intervention details (colchicine dosage, administration route, treatment duration), control group characteristics (standard of care, placebo, corticosteroid co-administration), and primary outcomes (mortality, ICU admissions, need for mechanical ventilation, hospitalization duration, and ICU stay length). Secondary outcomes included adverse events, BMI, and corticosteroid co-administration. A third reviewer cross-checked all extracted data to ensure accuracy, and any discrepancies were resolved by consensus.

### Risk of Bias Assessment

The Cochrane Risk of Bias Tool for Randomized Controlled Trials was used to assess the methodological quality of the included studies (Higgins et al., 2011). The tool evaluates seven domains: random sequence generation, allocation concealment, blinding of participants and personnel, blinding of outcome assessment, incomplete outcome data, selective reporting, and other potential sources of bias. Each study was classified as having low risk, some concerns, or a high risk of bias. Two independent reviewers conducted the risk assessment, with a third reviewer resolving any discrepancies.

### Data Synthesis and Statistical Analysis

Meta-analyses were conducted using R software with the *meta* and *dmetar* packages (Balduzzi et al., 2019; Harrer et al., 2019; Team, 2024). Binary outcomes, including mortality, ICU admission, and mechanical ventilation, were analyzed using risk differences (RD) with 95% Agresti-Coull confidence intervals (CI). Continuous outcomes, such as hospitalization duration and ICU stay, were assessed using mean differences (MD) with 95% CI. To account for expected variability among studies, a random-effects model employing the restricted maximum likelihood (REML) estimator was used. Heterogeneity was evaluated using the *I²* statistic.

For studies that did not directly report mean and standard deviation (SD) values but provided median and interquartile ranges (IQR), we applied conventional conversion methods to estimate missing SD values. For the non-primary outcome of age, two studies provided only IQRs, so the SD was estimated based on the 25th and 75th quantiles of the normal distribution. In one case where age SD was missing entirely, the 75th percentile of the SDs from other studies was used as a conservative estimate. Forest plots were generated to visualize effect sizes, allowing for a comprehensive assessment of colchicine’s clinical impact. To assess potential publication bias, we constructed funnel plots for all primary outcomes.

### Subgroup Analyses

To assess whether corticosteroid use modified colchicine’s effectiveness, we conducted subgroup analyses comparing studies that administered corticosteroids alongside colchicine to those that did not. Additionally, studies were stratified based on treatment duration (< 14 vs. > 14 days, < 21 vs. > 21 days, or 15 - 28 days) to determine whether colchicine’s efficacy varied with administration length. Further, differences in patient BMI and comorbidities were examined to assess whether specific patient characteristics influenced colchicine’s effectiveness.

## RESULTS

### Study Selection

A total of 249 articles were identified from PubMed (*n* = 76), EMBASE (*n* = 71), and Web of Science (*n* = 102). Six additional articles were retrieved from the references of systematic reviews. After removing 65 duplicates, 184 articles remained for screening. Two independent reviewers screened these articles, resulting in the exclusion of 168 that did not meet the inclusion criteria. Sixteen full-text articles were assessed for eligibility, of which three were excluded for not being randomized controlled trials (RCTs). 13 RCTs were included in the final systematic review and meta-analysis (Figure 2).

**Figure 2.**
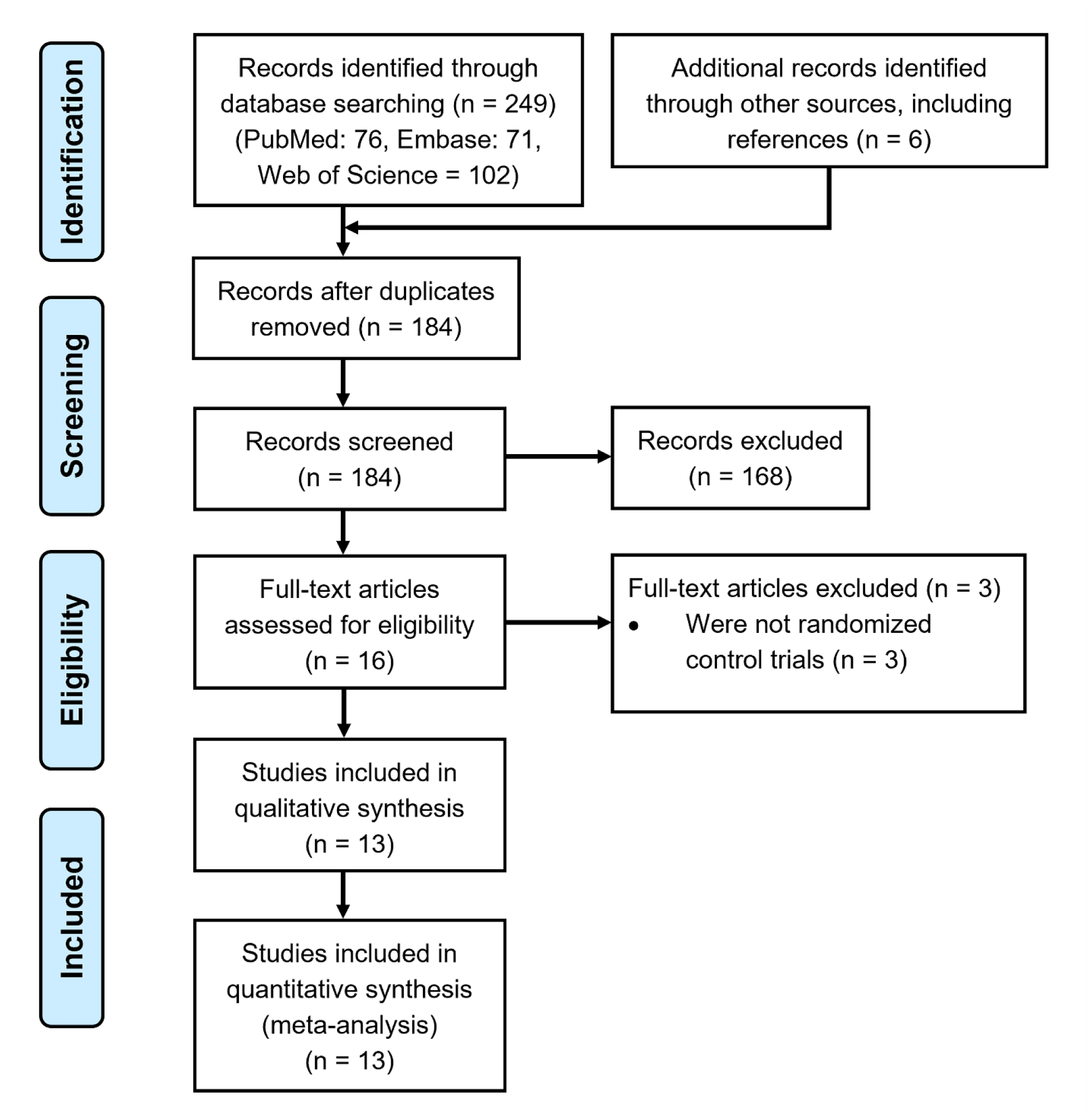
Flow diagram of study selection for the systematic review and meta-analysis on the efficacy of colchicine with or without corticosteroids in hospitalized COVID-19 patients. The diagram follows the Preferred Reporting Items for Systematic Reviews and Meta-Analyses (PRISMA) guidelines, detailing the identification, screening, eligibility, and inclusion process. A total of 249 records were identified through database searches (PubMed: 76, Embase: 71, Web of Science: 102), with an additional 6 records sourced from references. After removing duplicates, 184 records were screened, of which 168 were excluded. Sixteen full-text articles were assessed for eligibility, with 3 excluded for not being randomized controlled trials. Ultimately, 13 studies were included in both qualitative synthesis and quantitative meta-analysis. The flow diagram is adapted from the PRISMA approach (Moher et al., 2009).

### Study Characteristics

The 13 included studies involved a total of 16,529 participants, with 11,043 (66.8%) males and 5,486 (33.2%) females. The mean age across studies was 58.2 years (range: 46.1 – 69.1). Treatment regimens varied, with colchicine administered at loading doses of 0.5 mg to 2 mg, followed by maintenance doses ranging from 0.5 mg to 1 mg daily. Duration of treatment varied from 5 - 31 days. Control groups received SoC, which frequently included corticosteroids, anticoagulants, and antiviral treatments. Measurements of BMI were only recorded in five of the thirteen articles. Among the five articles, the overall mean was 26.4 kg/m with the lowest 22 kg/m and the highest 31.7kg/m (Table 1).

**Table 1.** Demographics of study participants in the selected randomized controlled trials.

| PMID | Reference | Treatment Group | No. of Participants | Sex |  | Mean | Age SD |
| --- | --- | --- | --- | --- | --- | --- | --- |
|  |  |  |  | # of Males | # of Females |  |  |
| 36302537 | Haroon et al., 2022 | colchicine | 48 | 31 | 17 | 53.5 | 17.4 |
|  |  | SoC<sup>1</sup> | 48 | 29 | 19 | 61.1 | 15.1 |
| 36396522 | Perricone et al., 2023 | colchicine | 77 | 47 | 30 | 69.1 | 13.1 |
|  |  | SoC | 75 | 50 | 25 | 67.9 | 15 |
| 36647298 | Kasiri et al., 2023 | colchicine | 55 | 27 | 28 | 53.2 | 15.28 |
|  |  | SoC | 51 | 21 | 30 | 56.16 | 11.71 |
| 36228641 | Eikelboom et al., 2022 | colchicine | 1304 | 762 | 542 | 56.1 | 16.7 |
|  |  | SoC | 1307 | 796 | 511 | 56 | 16 |
| 34672950 | RECOVERY, 2021 | colchicine | 5610 | 3897 | 1713 | 63.3 | 13.8 |
|  |  | SoC | 5730 | 4012 | 1718 | 63.5 | 13.7 |
| 33542047 | Lopes et al., 2021 | colchicine | 36 | 19 | 17 | 54.5<sup>2</sup> | (42.5-64.5)<sup>3</sup> |
|  |  | SoC | 36 | 14 | 22 | 55<sup>2</sup> | (42.0-67.0)<sup>3</sup> |
| 34964849 | Diaz et al., 2021 | colchicine | 640 | 421 | 219 | 62 | 14 |
|  |  | SoC | 639 | 409 | 230 | 62 | 15 |
| 34539185 | Pascual-Figal et al., 2021 | colchicine | 52 | 27 | 25 | 51.8 | 11.7 |
|  |  | SoC | 51 | 27 | 24 | 50.3 | 12.4 |
| 35654818 | Cecconi et al., 2022 | colchicine | 119 | 70 | 49 | 66 | 15.5 |
|  |  | SoC | 120 | 71 | 49 | 64.2 | 16.4 |
|  |  |  |  | # of Males | # of Females |  |  |
| 36128202 | Salehzadeh et al., 2022 | colchicine | 50 | 19 | 31 | 56.56 | 17.16 |
|  |  | SoC | 50 | 22 | 28 | 55.56 | 16.38 |
| 37075454 | Bonifácio et al., 2023 | colchicine | 14 | 10 | 4 | 56.2 | 11.6 |
|  |  | SoC | 16 | 11 | 5 | 46.1 | 10.6 |
| 32579195 | Deftereos et al., 2020 | colchicine | 55 | 31 | 24 | 63<sup>2</sup> | (55-70)<sup>3</sup> |
|  |  | SoC | 50 | 30 | 20 | 65<sup>2</sup> | (54-80)<sup>3</sup> |
| 36383611 | Rahman et al., 2022 | colchicine | 148 | 95 | 53 | 47<sup>4</sup> | (35-55)<sup>3</sup> |
|  |  | SoC | 148 | 95 | 53 |  |  |

<sup>1</sup> SoC – standard of care
<sup>2</sup> Median age
<sup>3</sup> Interquartile range for age
<sup>4</sup> Median age for the entire population, not specified between groups

### Treatment Regimen

The method, duration, and frequency of drug administration were recorded by all thirteen articles (Table 2). All thirteen studies included Colchicine and the placebo, when used, in oral form. Loading doses are common for many drug treatments due to the need for the achievement of proper drug concentrations at the beginning of any drug regimen. Nine studies included a larger loading dose of Colchicine before establishing a lower maintenance dose. The loading doses ranged in measurements between 0.5mg to 2mg, and all studies except one included a maintenance dose of 0.5-0.6mg, with the one being 1mg. Treatment frequency ranged between once to thrice daily; however, four studies changed frequency during the study’s duration, with the administration of the drug decreasing over time. Treatment duration ranged between studies, with 5 days as the shortest duration and 31 days as the longest duration. Of the thirteen studies, ten provided patients with corticosteroids in addition to Colchicine (Supplemental Table 4). Only one of these ten studies supported the use of Colchicine in their patient population. Three of the thirteen studies did not use any corticosteroids, and all three of these studies showed that Colchicine was effective in treating hospitalized COVID-19 patients.

**Table 2.** Baseline Characteristics and Clinical Outcomes of Included Studies.

| Reference<br>(PMID) | Treatment<br>Group | No. of<br>Study<br>Participants | Sex |  | Age |  | Treatment<br>Dose<sup>2</sup><br>(mg) | Treatment<br>Frequency<br>(times/day)<br>[No. of Days] | 30-Day<br>Mortality<br>Rate<br>(%) | Need for<br>Mechanical<br>ventilation<br>(%) |
| --- | --- | --- | --- | --- | --- | --- | --- | --- | --- | --- |
|  |  |  | # of<br>Males | # of<br>Females | Mean<br>(Median) | SD<br>(IQR)<sup>1</sup> |  |  |  |  |
| Haroon et al.,<br>2022<br>(36302537) | colchicine<br>SoC<sup>4</sup> | 48<br>48 | 31<br>29 | 17<br>19 | 53.5<br>61.1 | 17.4<br>15.1 | 0.5 | 1 to 2<sup>3</sup> [14] | 8.30<br>6.30 | -<br>- |
| Perricone et al.,<br>2023<br>(36396522) | colchicine<br>SoC | 77<br>75 | 47<br>50 | 30<br>25 | 69.1<br>67.9 | 13.1<br>15 | 0.5 to 1<sup>5</sup> | 2 to 3 [30] | 9.10<br>6.70 | 5.2<br>4 |
| Kasiri et al.,<br>2023<br>(36647298) | colchicine<br>SoC | 55<br>51 | 27<br>21 | 28<br>30 | 53.2<br>56.16 | 15.28<br>11.71 | 0.5 | 2 [7] | 10.90<br>11.80 | -<br>- |
| Eikelboom et al.,<br>2022<br>(36228641) | colchicine<br>SoC | 1304<br>1307 | 762<br>796 | 542<br>511 | 56.1<br>56 | 16.7<br>16 | 0.6 | 2 [28] | 20.20<br>19.10 | 18.9<br>19.3 |
| RECOVERY,<br>2021<br>(34672950) | colchicine<br>SoC | 5610<br>5730 | 3897<br>4012 | 1713<br>1718 | 63.3<br>63.5 | 13.8<br>13.7 | 0.5 | 2 [10] | 21<br>21 | 11<br>11 |
| Lopes et al.,<br>2021<br>(33542047) | colchicine<br>SoC | 36<br>36 | 19<br>14 | 17<br>22 | (54.5)<br>(55) | (42.5-64.5)<br>(42.0-67.0) | 0.5 | 2 to 3<sup>6</sup> [5] | 0<br>5.56 | -<br>- |
| Diaz et al., 2021<br>(34964849) | colchicine<br>SoC | 640<br>639 | 421<br>409 | 219<br>230 | 62<br>62 | 14<br>15 | 0.5 | 2 [14] | 20.50<br>22.20 | 4.5<br>6.6 |
| Pascual-Figal et<br>al., 2021<br>(34539185) | colchicine<br>SoC | 52<br>51 | 27<br>27 | 25<br>24 | 51.8<br>50.3 | 11.7<br>12.4 | 0.5 | 1 to 2<sup>7</sup> [28] | 0<br>3.90 | 0<br>3.9 |
|  |  |  | # of<br>Males | # of<br>Females | Mean<br>(Median) | SD<br>(IQR)<sup>1</sup> |  |  |  |  |
| Cecconi et al.,<br>2022<br>(35654818) | colchicine<br>SoC | 119<br>120 | 70<br>71 | 49<br>49 | 66<br>64.2 | 15.5<br>16.4 | 0.5 | 1 [5] | 5.90<br>8.33 | 4.2<br>8.3 |
| Salehzadeh et<br>al., 2022<br>(36128202) | colchicine<br>SoC | 50<br>50 | 19<br>22 | 31<br>28 | 56.56<br>55.56 | 17.16<br>16.38 | 1 | 1 [6] | -<br>- | -<br>- |
| Bonifácio et al.,<br>2023<br>(37075454) | colchicine<br>SoC | 14<br>16 | 10<br>11 | 4<br>5 | 56.2<br>46.1 | 11.6<br>10.6 | 0.5 | 2 [28] | 0<br>12.50 | -<br>- |
| Deftereos et al.,<br>2020<br>(32579195) | colchicine<br>SoC | 55<br>50 | 31<br>30 | 24<br>20 | (63)<br>(65) | (55-70)<br>(54-80) | 0.5 | 2 [21] | 3<sup>8</sup><br>17<sup>8</sup> | -<br>- |
| Rahman et al.,<br>2022<br>(36383611) | colchicine<br>SoC | 148<br>148 | 95<br>95 | 53<br>53 | (47)<sup>9</sup> | (35-55) | 0.6 | 1 [28] | 2.7<br>8.9 | 1.4<br>2.7 |

<sup>1</sup> Interquartile range for age
<sup>2</sup> Maintenance dose
<sup>3</sup> First six days, the drug was given twice daily and the second week it was given once a day
<sup>4</sup> SoC – standard of care
<sup>5</sup> 0.5mg if weight was <100kg or 1mg if weight was >100kg
<sup>6</sup> First five days, colchicine was given three times daily, and then for the second five days, colchicine was given twice daily
<sup>7</sup> 0.5mg twice a day for one week and 0.5mg once per day for 28 days
<sup>8</sup> 21-Day mortality
<sup>9</sup> Median age for the entire population, not specified between groups

### Risk of Bias Assessment

Using the Cochrane Collaboration tool for risk of bias assessment (Table 3), the estimated bias introduced in each of the thirteen studies was relatively low between the seven categories. While some studies demonstrated a well-controlled design, others presented some limitations. Only one study was recorded as having a high risk of bias in three categories, which included allocation concealment, blinding of participants and personnel, and blinding of outcome assessment (Eikelboom et al., 2022). Four additional studies (Bonifácio et al., 2023; Deftereos et al., 2020; Haroon et al., 2022; Pascual-Figal et al., 2021) exhibited high risk of bias in two categories, while two studies (Lopes et al., 2021; Perricone et al., 2023) had a mix of high risk and some concern ratings, suggesting potential methodological weaknesses. Allocation concealment contained the most high-risk markings, with eight of the thirteen articles failing this assessment. This increases the likelihood of selection bias, which may influence treatment outcomes. It should be noted that open-label drug trials were common within the first year of the COVID-19 pandemic, introducing potential performance and detection bias.

**Table 3.** Risk of bias assessment of randomized controlled trial (RCT) studies using the Cochrane Collaboration’s tool for assessing risk of bias in randomized trials.

| Reference | PMID | Random<br>sequence<br>generation | Allocation<br>concealment | Blinding of<br>participants<br>and<br>personnel | Blinding of<br>outcome<br>assessment | Incomplete<br>outcome<br>data | Selective<br>reporting | Other<br>sources of<br>bias<sup>1</sup> |
| --- | --- | --- | --- | --- | --- | --- | --- | --- |
| Haroon et al., 2022 | 36302537 | + | - | - | ? | + | + | ? |
| Perricone et al., 2023 | 36396522 | + | - | ? | ? | - | + | + |
| Kasiri et al., 2023 | 36647298 | + | + | + | + | + | + | ? |
| Eikelboom et al., 2022 | 36228641 | + | - | - | - | + | + | + |
| RECOVERY, 2021 | 34672950 | + | - | - | + | ? | + | + |
| Lopes et al., 2021 | 33542047 | + | + | + | ? | - | + | ? |
| Diaz et al., 2021 | 34964849 | + | - | ? | + | + | + | ? |
| Pascual-Figal et al.,<br>2021 | 34539185 | + | - | - | ? | + | + | + |
| Cecconi et al., 2022 | 35654818 | + | + | ? | + | + | + | ? |
| Salehzadeh et al., 2022 | 36128202 | ? | + | + | ? | + | ? | + |
| Bonifácio et al., 2023 | 37075454 | + | - | - | ? | + | + | + |
| Deftereos et al., 2020 | 32579195 | ? | - | - | ? | + | + | ? |
| Rahman et al., 2022 | 36383611 | + | + | + | + | + | + | ? |

Key: + Low risk of bias, - High risk of bias, ? Some concern of bias

### Meta-Analysis of Primary Outcomes

For overall mortality, colchicine was associated with a 2% absolute reduction (RD = −0.02, 95% CI: [-0.04, 0.00]), but this result did not reach statistical significance (Figure 3a). The moderate heterogeneity (I² = 38%) suggests some variability among the included studies, potentially due to differences in patient populations, study designs, and background treatments. For mechanical ventilation, colchicine showed a small reduction in MV rates (RD = −0.01, 95% CI: [-0.02, 0.00]) with low heterogeneity (I² = 8%) (Figure 3c).

**Figure 3.**
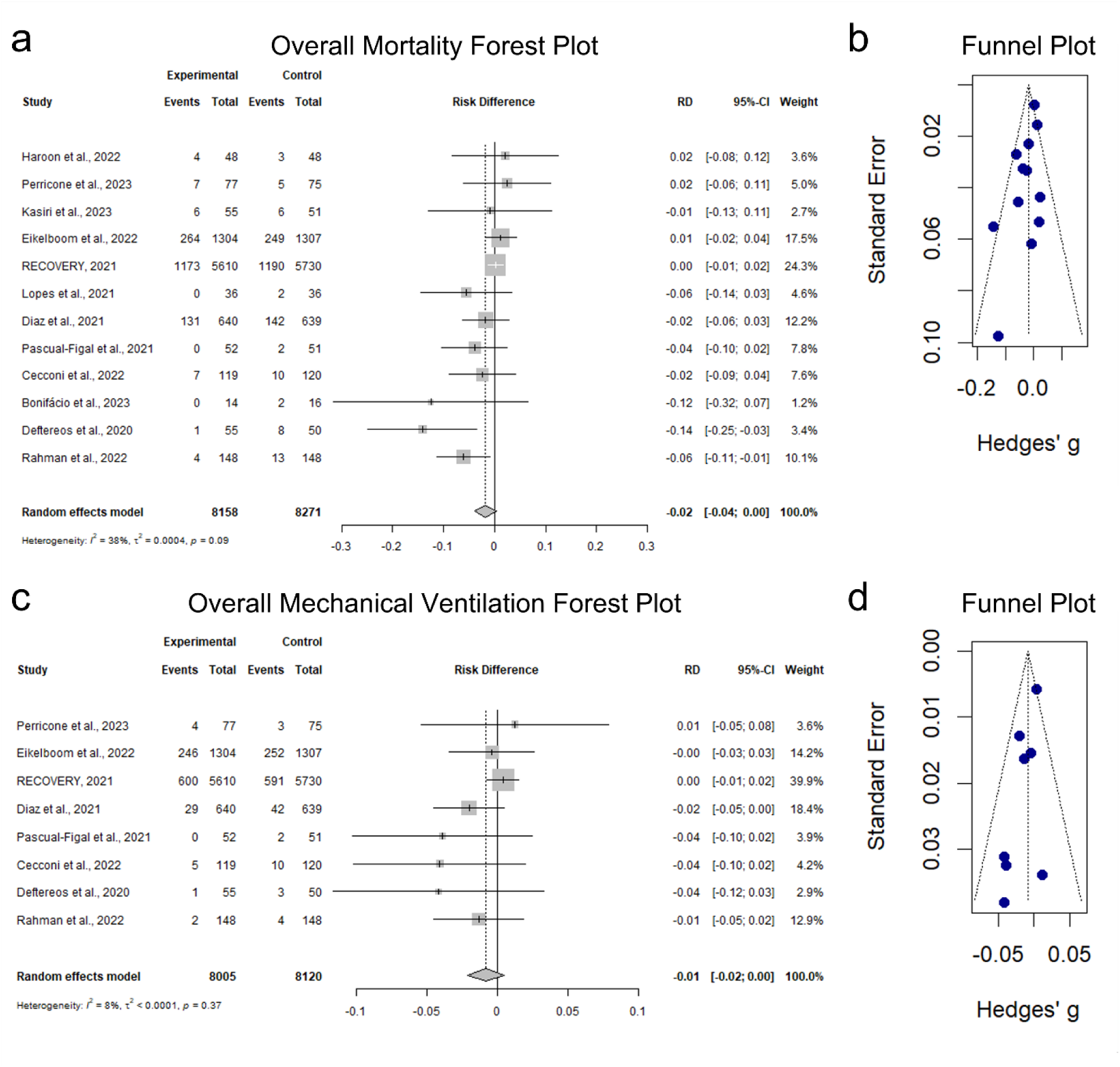
Meta-analysis results for the efficacy of colchicine in hospitalized COVID-19 patients. (a) Forest plot showing the risk difference (RD) in overall mortality between patients receiving colchicine (n = 8,158) and those not receiving colchicine (n = 8,271). The pooled RD is −0.02 (95% CI: [-0.04, 0.00]), with a heterogeneity index (I²) of 38%. (b) Funnel plot assessing publication bias for studies reporting overall mortality. (c) Forest plot showing the risk difference (RD) in overall mechanical ventilation between patients receiving colchicine (n = 8,005) and those not receiving colchicine (n = 8,120). The pooled RD is −0.01 (95% CI: [-0.02, 0.00]), with a heterogeneity index (I²) of 8%. (d) Funnel plot assessing publication bias for studies reporting mechanical ventilation outcomes.

When examining ICU admissions, colchicine was associated with a 3% absolute reduction (RD = −0.03, 95% CI: [-0.06, 0.01]), but this effect was not statistically significant (Supplemental Figure 1a). Notably, heterogeneity was absent (I² = 0%). For hospitalization duration, colchicine did not demonstrate a significant reduction (MD = −0.58 days, 95% CI: [-1.80, 0.64]) and had high heterogeneity (I² = 68%) (Supplemental Figure 1c).

### Impact of Corticosteroids on Colchicine’s Effect

A key finding was the association between colchicine and corticosteroids. When stratifying the analysis by corticosteroid use, results indicated that colchicine was significantly associated with a 7% reduction in mortality among patients who did not receive corticosteroids (RD = −0.07, 95% CI: [−0.11, −0.03]) (Figure 4a). However, for patients who received corticosteroids as part of their standard of care, colchicine did not affect mortality (RD = 0, 95% CI: [-0.01, 0.01]). This suggests that corticosteroids may diminish colchicine’s potential benefits, possibly due to overlapping anti-inflammatory mechanisms that make colchicine redundant in patients already receiving potent corticosteroid therapy.

**Figure 4.**
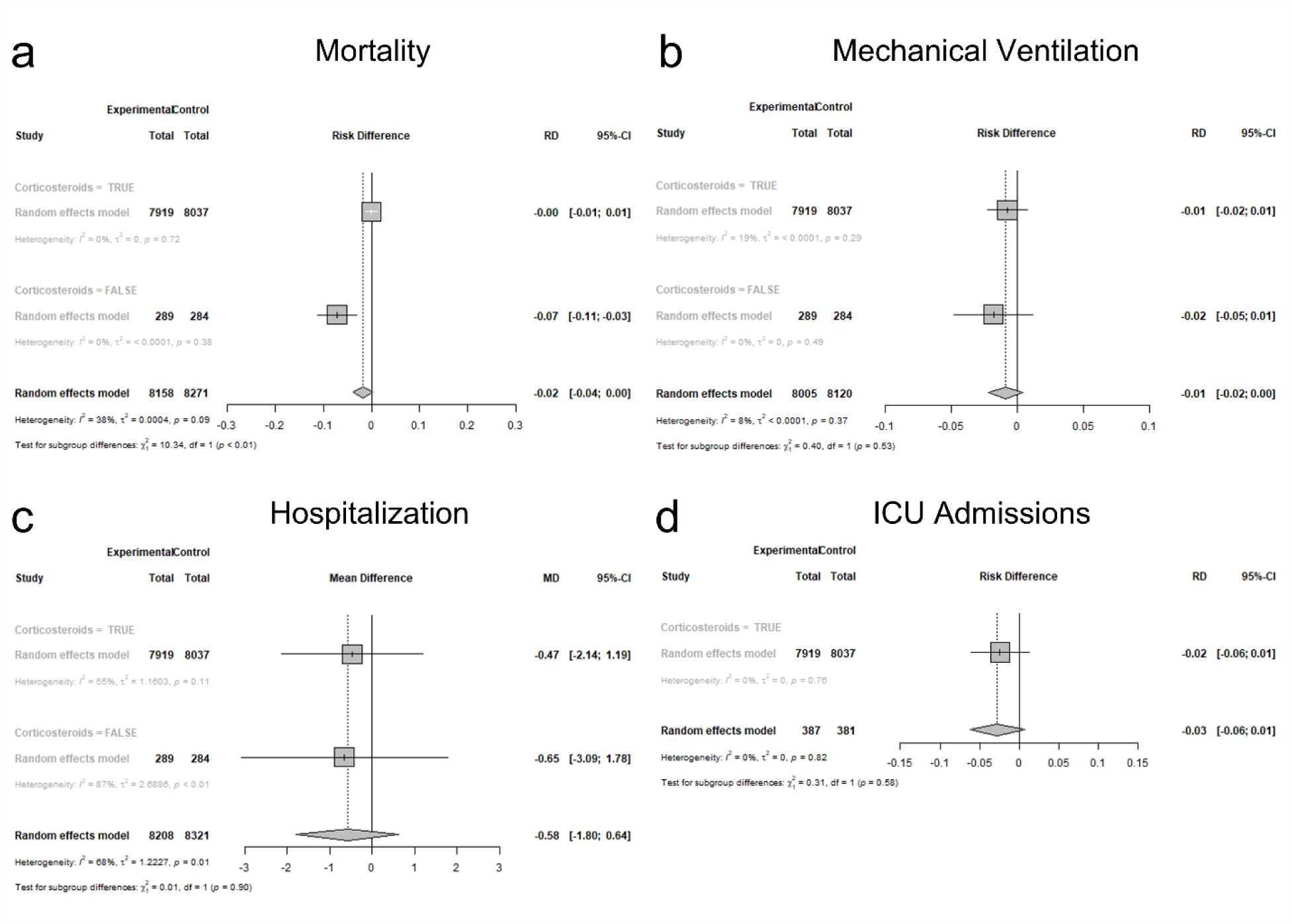
Subgroup meta-analysis of colchicine efficacy in hospitalized COVID-19 patients with and without corticosteroids in hospitalized COVID-19 patients. (a) Forest plot showing the risk difference (RD) for mortality in patients receiving colchicine with corticosteroids (n = 7,919) and without corticosteroids (n = 289) compared to respective control groups. The RD for mortality with corticosteroids was 0.00 (95% CI: [-0.01, 0.01], I² = 0%), while the RD without corticosteroids was −0.07 (95% CI: [-0.11, −0.03], I² = 0%). (b) Forest plot of mechanical ventilation (MV) risk difference in colchicine-treated patients with corticosteroids (n = 7,919) and without corticosteroids (n = 289) compared to respective controls. The RD for MV with corticosteroids was −0.01 (95% CI: [-0.02, 0.01], I² = 19%), and the RD without corticosteroids was −0.02 (95% CI: [-0.05, 0.01], I² = 0%). (c) Forest plot of hospitalization risk difference in colchicine-treated patients with corticosteroids (n = 7,919) and without corticosteroids (n = 289) compared to respective controls. The RD for hospitalization with corticosteroids was −0.47 (95% CI: [-2.14, 1.19], I² = 55%), while the RD without corticosteroids was −0.65 (95% CI: [-3.09, 1.78], I² = 87%). (d) Forest plot of intensive care unit (ICU) admissions risk difference in colchicine-treated patients with corticosteroids (n = 7,919) and without corticosteroids (n = 387) compared to respective controls. The RD for ICU admissions with corticosteroids was −0.02 (95% CI: [-0.06, 0.01], I² = 0%), and the RD without corticosteroids was −0.03 (95% CI: [-0.06, 0.01], I² = 0%).

For hospitalization duration, colchicine was not associated with a significant reduction in either corticosteroid subgroup, and heterogeneity was high (I² = 55% with corticosteroids and I² = 87% without corticosteroids, Figure 4c). Similarly, ICU admissions and mechanical ventilation outcomes showed no statistically significant differences based on corticosteroid use, further reinforcing the hypothesis that colchicine’s effectiveness may be masked when corticosteroids are present.

### Impact of Treatment Duration on Colchicine’s Effect

For hospitalization duration, colchicine was associated with a significant reduction in hospital stay when administered for less than 21 days (MD = −1.77 days, 95% CI: [−2.61, −0.93], I² = 0%, Supplemental Figure 5b), but not for longer durations, suggesting that effectiveness of colchicine may only reside in shorter treatment courses.

### Assessment of Heterogeneity Among Study Results

High heterogeneity was observed for hospitalization duration (>14 days, I² = 51%) and mortality (>21 days, I² = 64%), suggesting that differences in patient populations, treatment protocols, and study designs contributed to variability in outcomes. Conversely, ICU admissions and mechanical ventilation outcomes had low to absent heterogeneity (I² = 0 - 8%), indicating greater consistency across studies (Supplemental Figure 2). To explore potential sources of heterogeneity, subgroup analyses were conducted based on corticosteroid use and treatment duration. These analyses aimed to identify whether colchicine’s effects varied depending on associated anti-inflammatory treatments or duration of therapy.

A key finding was that studies where corticosteroids were not included in the standard of care were more likely to report a statistically significant benefit of colchicine. Conversely, studies that administered colchicine alongside dexamethasone or other corticosteroids generally found no significant differences between treatment and control groups, suggesting a potential interaction between these two anti-inflammatory agents. Another important factor influencing heterogeneity was BMI. Some studies indicated that patients with higher BMI had improved outcomes with colchicine treatment. However, due to the limited number of studies reporting BMI data, the extent to which body mass influences colchicine’s efficacy remains uncertain and requires further research.

## Conclusions

In the overall population of hospitalized COVID-19 patients, colchicine did not significantly improve mortality, ICU admissions, mechanical ventilation rates, or ICU stay. However, in patients who did not receive corticosteroids, colchicine significantly reduced mortality by 7%, suggesting a potential benefit in this subgroup. Additionally, colchicine significantly reduced hospitalization duration when used for less than 21 days. These findings highlight the potential clinical utility of colchicine in specific patient populations, particularly those not receiving corticosteroids and those requiring shorter hospital stays. Further research should focus on identifying optimal patient subgroups and treatment durations to maximize colchicine’s therapeutic potential in COVID-19 management.

## DISCUSSION

Among the thirteen articles selected and qualitatively analyzed, the overall support for colchicine against COVID-19 was not found due to the lack of statistical significance reported by most trials. When reviewing the data within the five outcome categories, many studies recorded the treatment group experiencing some improvement compared to their control group counterparts. However, most of these data sets were not statistically significant and, therefore, not emphasized by the authors.

Corticosteroids have been widely studied as a treatment for severe COVID-19, particularly in hospitalized patients requiring respiratory support. A recent systematic review and meta-analysis found that corticosteroid therapy significantly improved survival rates (Patel et al., 2022). However, low-dose corticosteroids did not show a clear benefit, emphasizing the importance of appropriate dosing. Another systematic review through July 2020 found that hospitalized COVID-19 patients in the ICU or on mechanical ventilation had higher survival rates when treated with corticosteroids (van Paassen et al., 2020). These findings reinforce the crucial role of corticosteroids in severe COVID-19 management, particularly in patients with hyperinflammation and respiratory failure, while highlighting the need for optimized dosing strategies to maximize benefits and minimize risks.

The influence of corticosteroids among colchicine recipients should be considered, as both drugs act as anti-inflammatory agents within the immune system. Of the four studies that found colchicine to be effective against COVID-19, three did not report corticosteroid use within the standard care treatment plan. The possibility of corticosteroids such as dexamethasone reducing colchicine’s strength or interfering with the drug’s pathway should be considered in further research. When considering the STIMULATE ICP trial and proper treatment for long COVID symptoms, colchicine has the potential to be a distinguished drug of choice for reducing chronic inflammation if dexamethasone or other corticosteroids are not present.

The findings of this review align with some previous meta-analyses that did not find a significant benefit of colchicine in reducing mortality among hospitalized COVID-19 patients. However, this study uniquely identifies corticosteroid use as a key factor in colchicine’s efficacy. Unlike previous reviews, which broadly assessed colchicine’s impact, our findings suggest that colchicine might only be effective in the absence of corticosteroids. This aligns with mechanistic studies suggesting that both colchicine and corticosteroids target inflammation through overlapping pathways, potentially rendering colchicine redundant when corticosteroids are present (Bonaventura et al., 2022). Future studies should further explore this interaction in controlled settings.

The studies included in this meta-analysis varied in their methodology, sample sizes, and treatment regimens, leading to substantial heterogeneity in some analyses (e.g., hospitalization duration, I² = 68%). Additionally, the risk of bias was a concern in several studies, particularly regarding allocation concealment (high risk in 8 studies) and open-label designs. The RECOVERY trial, which contributed a large proportion of the total sample size (*n* = 11,340), heavily influenced the pooled effect estimates. While this strengthens statistical power, it also means that the overall results may be disproportionately reflective of this single large trial rather than a balanced synthesis of all available evidence. Future research should aim for well-controlled trials with standardized protocols to reduce these limitations. This review included a comprehensive meta-analysis of 13 randomized controlled trials, synthesizing findings across 16,529 patients. While every effort was made to include all relevant studies, publication bias remains a potential limitation. Funnel plots (Figure 3b) suggest a slight asymmetry, indicating a possible underrepresentation of smaller studies with negative findings.

Overall, this study concludes that though colchicine was not found to be an efficacious treatment against hospitalized COVID-19 patients, certain factors contributed to its unfavorable results. One such factor was the frequent use of corticosteroids alongside colchicine in treating hospitalized COVID-19 patients during the early stages of the pandemic. Therefore, this did not provide a proper consideration for the efficacy of colchicine in treating COVID-19 patients. Future randomized controlled trials should focus on stratifying patients based on corticosteroid use and BMI to further elucidate these interactions. Additionally, given emerging concerns about long COVID, future studies should investigate colchicine’s potential role in managing chronic inflammation in post-acute COVID-19 syndromes.

## Funding

This research received no specific grant from any funding agency.

## Conflict of Interest

The authors declare no conflicts of interest relevant to the content of this manuscript.

## CRediT Author Statement

**Marisa Arthur**: Conceptualization; Investigation; Data Curation; Visualization; Writing – Original Draft; Writing – Review & Editing

**Jason Wilson**: Formal Analysis; Software; Visualization; Writing – Original Draft; Writing – Review & Editing; Supervision

**Enze Cai**: Formal Analysis

**JongWon See**: Investigation

**Brent M. Peterson**: Investigation; Writing – Review & Editing

**Thushara Galbadage**: Conceptualization; Methodology; Investigation; Data Curation; Writing – Original Draft; Writing – Review & Editing; Supervision; Project Administration

## Data Availability

All data produced in the present study are available upon reasonable request to the authors.

## Notes

### Competing Interest Statement

The authors have declared no competing interest.

### Clinical Protocols

https://www.crd.york.ac.uk/PROSPERO/view/CRD42024510295

### Funding Statement

This study did not receive any funding.

