## Supplemental Information for "Efficacy of colchicine with or without corticosteroids in hospitalized COVID-19 patients: A systematic review and meta-analysis"

Marisa Arthur[1], Jason Wilson[2], Enze Cai[3], JongWon See[1], Brent M. Peterson[1], Thushara Galbadage[1*#]

[1] Department of Kinesiology and Public Health, Biola University, La Mirada, CA

[2] Department of Mathematics and Computer Science, Biola University, La Mirada, CA

[3] Department of Statistics, University of Illinois Urbana-Champaign, Champaign, IL

[#] Now in the Department of Applied Health Sciences, Texas Christian University, Fort Worth, TX, US

**Keywords:** Intervention, clinical, cytokine storm, anti-inflammatory, mortality, hospitalization, corticosteroids, randomized controlled trial (RCT), long COVID

**Supplemental Table 1**. Previous systematic reviews and meta-analyses studying the efficacy of colchicine in hospitalized COVID-19 patients.

| PMID | Reference | Year[1] | RCT only[2] | Supported the use of colchicine |
|---|---|---|---|---|
| 34658014 | Mikolajewska et al., 2021 | 2021 | Yes | No |
| 35566737 | Toro-Huamanchumo et al., 2022 | 2022 | No | No |
| 34440608 | Lien et al., 2021 | 2021 | No | Yes[3] |
| 35258357 | Sanghavi et al., 2022 | 2022 | No | No |
| 34962939 | Chiu et al., 2021 | 2021 | No | Yes[3] |
| 33617817 | Salah et al., 2021 | 2021 | No | Yes[3] |
| 35078098 | Zein et al., 2022 | 2022 | No | No |
| 33719081 | Hariyanto et al., 2021 | 2021 | No | Yes |
| 35833737 | Lan et al., 2022 | 2022 | Yes | No |
| 34185313 | Elshafei et al., 2021 | 2021 | No | Yes[3] |
| 35381033 | Yasmin et al., 2022 | 2022 | Yes | Yes |
| 33421583 | Vrachatis et al., 2021 | 2021 | No | Yes[3] |
| 34162130 | Nawangsih et al., 2021 | 2021 | No | Yes[3] |
| 34250834 | Golpour et al., 2021 | 2021 | No | Yes |
| 34970856 | Kow et al., 2022 | 2022 | Yes | Yes[4] |
| 35446263 | Romeo et al., 2022 | 2022 | No | No |
| 34859881 | De-Miguel-Balsa et al., 2021 | 2021 | No | No |
| 34810227 | Mehta et al., 2021 | 2021 | Yes | No |

[1] Year of publication

[2] The systematic reviews and meta-analysis included randomized clinical trials only

[3] Supported the use of colchicine for mortality reduction benefits

[4] Supported the use of colchicine for reduced hospitalization but not for mortality benefits

**Supplemental Table 2.** Randomized control trials (RCTs) used in this systematic review and meta-analysis.

| PMID | Reference | Clinical Trial Registration # | Study End Date | Times Cited[1] |
|---|---|---|---|---|
| 33542047 | (Lopes et al., 2021) | RBR-8jyhxh[4] | 08/30/2020 | 18 |
| 32579195 | (Deftereos et al., 2020) | Not Available[6] | 04/27/2020 | 18 |
| 34672950 | (RECOVERY, 2021) | NCT04381936 | 03/04/2021 | 10 |
| 36128202 | (Salehzadeh et al., 2022) | IRCT20200418047126N1[3] | 06/20/2020 | 5 |
| 34964849 | (Diaz et al., 2021) | NCT04328480 | 04/28/2021 | 3 |
| 34539185 | (Pascual-Figal et al., 2021) | NCT04350320 | 12/04/2020 | 3 |
| 36302537 | (Haroon et al., 2022) | NCT04667780[2] | 07/07/2021 | 0 |
| 36396522 | (Perricone et al., 2023) | NCT04375202 | 05/12/2021 | 0 |
| 36647298 | (Kasiri et al., 2023) | IRCT20190804044429N5[3] | 05/1/2021 | 0 |
| 36228641 | (Eikelboom et al., 2022) | NCT04324463 | 02/10/2022 | 0 |
| 35654818 | (Cecconi et al., 2022) | EUCTR2020-001841-38-ES [5] | 03/1/2021 | 0 |
| 37075454 | (Bonifácio et al., 2023) | NCT04724629 | 06/09/2021 | 0 |
| 36383611 | (Rahman et al., 2022) | NCT04527562 | 11/15/2020 | 0 |

[1] Number of times cited in previous systematic reviews and meta-analyses.
[2] NCT articles are registered through the clinicaltrials.org
[3] Registered through the Iranian Registry of Clinical Trials
[4] Registered through the National Registry, Brazil
[5] Registered and approved by the EU (European Union) Clinical Trials Register
[6] Registered through the National Ethics Committee and the Hellenic National Organization for Medicines

**Supplemental Table 3.** Literature Search Strategy and Results

| Database | Search Query | Limits Applied | Results |
|---|---|---|---|
| PubMed | (colchicine) AND (COVID OR COVID-19 OR SARS-CoV-2) AND (hospitalized OR hospital) AND (clinical trial OR randomized controlled trial OR RCT) | None | 76 |
| Embase | (colchicine) AND (COVID OR COVID-19 OR SARS-CoV-2) AND (hospitalized OR hospital) AND (clinical trial OR randomized controlled trial OR RCT) | Controlled Clinical Trial, Randomized Controlled Trial | 71 |
| Web of Science | (colchicine) AND (COVID OR COVID-19 OR SARS-CoV-2) AND (hospitalized OR hospital) AND (clinical trial OR randomized controlled trial OR RCT) | None | 102 |

**Supplemental Table 4.** Observed Trends

| PMID | Reference | Treatment Duration | Used Corticosteroids[1] | Overall Support for Colchicine |
|---|---|---|---|---|
| 36302537 | Haroon et al., 2022 | 6 | Yes | No |
| 36396522 | Perricone et al., 2023 | 30 | Yes | No |
| 36647298 | Kasiri et al., 2023 | 7 | Yes | No |
| 36228641 | Eikelboom et al., 2022 | 28 | Yes | No |
| 34672950 | RECOVERY, 2021 | 10 | Yes | No |
| 33542047 | Lopes et al., 2021 | 10 | No | Yes |
| 34964849 | Diaz et al., 2021 | 14 | Yes | No |
| 34539185 | Pascual-Figal et al., 2021 | 28 | Yes | No |
| 35654818 | Cecconi et al., 2022 | 5 | Yes | No |
| 36128202 | Salehzadeh et al., 2022 | 6 | No | Yes |
| 37075454 | Bonifácio et al., 2023 | 31 | Yes | Yes |
| 32579195 | Deftereos et al., 2020 | 21 | No | Yes |
| 36383611 | Rahman et al., 2022 | 13 | Yes | No |

[1]Dexamethasone was the most commonly used corticosteroid

**Supplemental Table 5.** Overall summary of each article

| PMID | Reference | Summaries |
|---|---|---|
| 36302537 | Haroon et al., 2022 | - Two hospitalized non-ICU groups were studied in this treatment plan<br>- Those in the colchicine group were 7.6 years younger on average than the control group and had less comorbidities than the controls<br>- Colchicine was added to SOC which included high doses of corticosteroids<br>- The hypothesis was not supported as colchicine failed to show any significant improvement in hospitalization stay, ICU admission, and mortality levels |
| 36396522 | Perricone et al., 2023 | - Two groups of hospitalized non-ICU patients were split<br>- Both received standard of care and the treatment group included colchicine<br>- This study did not find any difference in treatment efficacy in the colchicine group compared to the SOC group<br>- ICU admission was more prominent in the control group (4 individuals) than the colchicine group (1) but this was not statistically significant<br>- This experiment was conducted in the first wave of the pandemic and no loading dose was introduced |
| 36647298 | Kasiri et al., 2023 | - Hospitalized patients were divided into treatment and control groups.<br>- The loading dose was high and maintained by a lower daily dose for 7 days<br>- No significance was found between the outcomes of both groups<br>- Limitations included a lack of PCR tests administered and no studies on inflammatory cytokines |
| 36228641 | Eikelboom et al., 2022 | - A large open-label trial was conducted with 2000+ patients positive for COVID-19, hospitalized, and non-ICU<br>- Colchicine was given alongside SOC (usual care)<br>- There was no significant improvement in respiratory ventilation and mortality.<br>- This RCT had a high adherence rate but was tested over a long period of time (thus, introducing potential new variants) |

| | | |
|---|---|---|
| 34672950 | RECOVERY, 2021 | - A large well-performed RCT included 11,000 COVID-19 hospitalized patients<br>- Those who received Colchicine did not report significant improvement in ventilation, mortality, and duration of hospitalization stay.<br>- It was open-label, and no radiological levels were collected |
| 33542047 | Lopes et al., 2021 | - Just over 100 patients were enrolled in this study and randomized into two groups<br>- Both groups received SOC<br>- There was a significant difference in favor of the colchicine group for the use of supplemental oxygen (with a shorter need) and hospitalization stay.<br>- CRP levels were greatly reduced in the colchicine group at the beginning of treatment and LDH levels throughout the trial<br>- This trial was double-blinded, the participants were mainly females with high BMIs and used the maximum safe daily dose of the drug (considering a body weight of 50 kg, that is, 0.030 mg/kg). Colchicine was reported as effective |
| 34964849 | Diaz et al., 2021 | - A year-long trial was conducted with over 500 patients in each group<br>- Half of the patients had pre-existing hypertension<br>- 91% of patients received corticosteroids alongside SOC in both groups<br>- There were slight differences in favor of the treatment group for 28-day mortality and the need for mechanical invasive ventilation, but it was not statistically significant<br>- It should be noted that it was open-labeled. |
| 34539185 | Pascual-Figal et al., 2021 | - 100 patients were recruited in this trial<br>- 27% were diabetic and 21% were obese<br>- SOC was given to both groups including dexamethasone/heparin and remdesivir (not for all patients but occurred more in the treatment group)<br>- Clinical deterioration was in favor of the control group while ICU admission/mechanical ventilation was in favor of colchicine but was not significant.<br>- Colchicine did help to avoid further deterioration |

| | | |
|---|---|---|
| | | - Colchicine emerged as a significant protective factor for 1-point deterioration on the WHO scale for clinical improvement |
| 35654818 | Cecconi et al., 2022 | - Two groups of hospitalized patients were randomly divided into two groups<br>- Mean BMI was 27.5 (+- 4.5)<br>- Almost all patients received steroids (dexamethasone) and heparin<br>- Outcomes between the two groups were similar; however, the overweight patients in the treatment group showed a lower prevalence in the primary outcomes.<br>- Higher levels of ferritin (inflammation) were found in the colchicine group when looking at inflammatory markers T<br>- There was a noticeable improvement for the patients with a higher BMI. |
| 36128202 | Salehzadeh et al., 2022 | - 100 patients were randomly assigned to the two groups<br>- 59% of the patients were female and COPD was the most prevalent disease among the patients<br>- A significant difference (in favor of the colchicine group) regarding the time of fever and duration of hospitalization. |
| 37075454 | Bonifácio et al., 2023 | - A group of sixty patients were assigned to the four groups (one being SOC as the control). 14 patients were enrolled in the colchicine group<br>- Standard of Care included supplemental O2 ventilation, corticosteroids, anticoagulants, and/or antibiotics (if needed).<br>- Among the 60, 61% were male and the mean BMI was 31.7 kg/m. Improvement in the WHO scale occurred in all 14 patients of the colchicine group was 100% and none presented clinical deterioration.<br>- These findings were not statistically significant |
| 32579195 | Deftereos et al., 2020 | - A little over 100 patients were recruited for this study<br>- All patients in both groups received the following drugs in the SOC (chloroquine or hydroxychloroquine and azithromycin).<br>- When assessing outcomes that included the need for mechanical ventilation and mortality levels, all but 1 in the colchicine group did not meet the clinical primary endpoint (unlike 7 in the control) and eventually died |

| | | |
|---|---|---|
| | | - D-dimer levels were significantly lower in the treatment group |
| 36383611 | Rahman et al., 2022 | - 296 patients were randomly divided into treatment and control groups (the control being SOC which included paracetamol, antihistamines, and oxygen therapy)<br>- No difference in clinical deterioration was observed<br>- Colchicine reduced mortality and the need for mechanical ventilation, but these data points were not significant (56% reduction)<br>-<br>- The 28-day follow-up found a 2-point deterioration in 13 of the control groups and only 4 in the colchicine groups (significantly lower)<br>- Limitations including the short duration of treatment, the bitterness of the colchicine tablet, and the follow-up included patients no longer hospitalized |

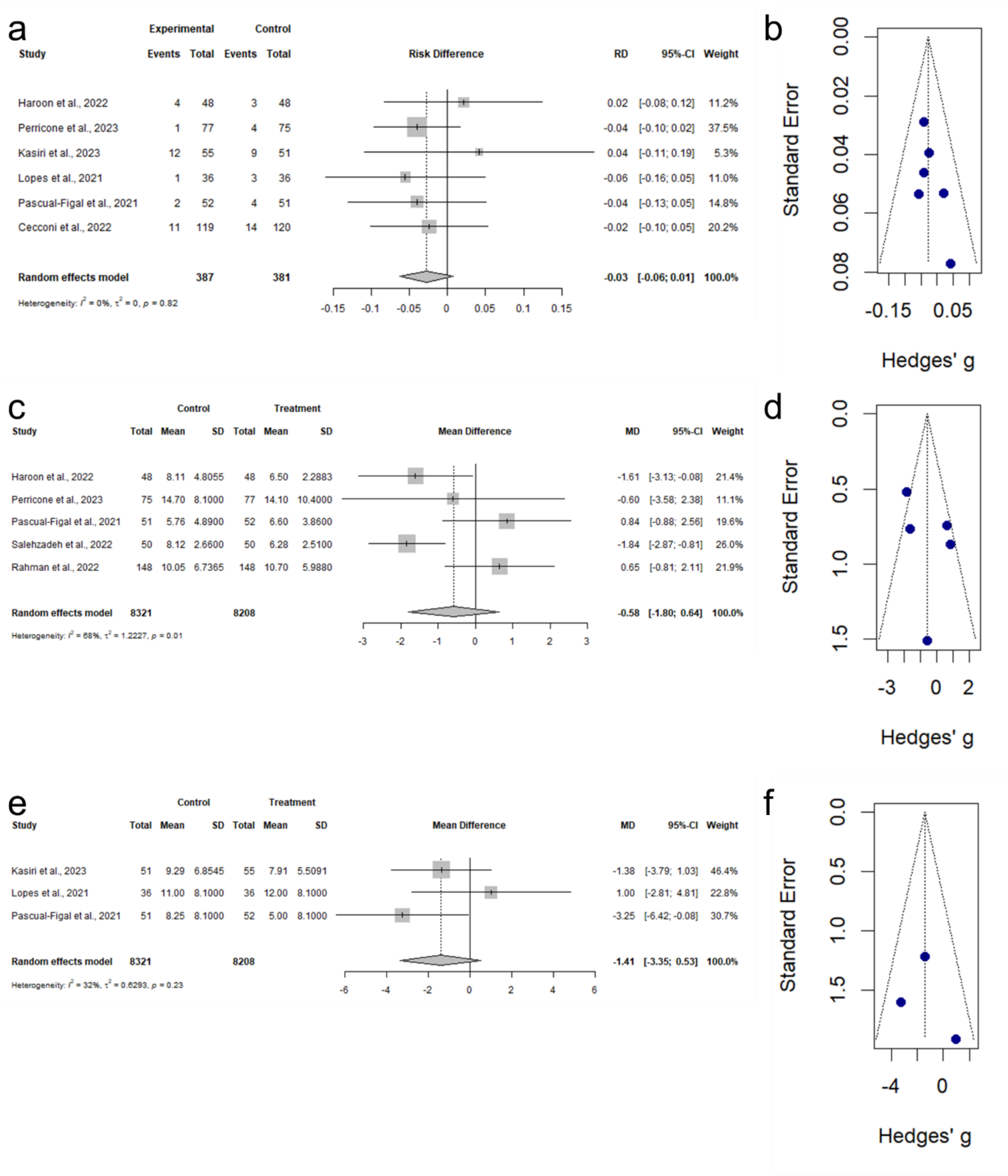


**Supplemental Figure 01. Overall efficacy of colchicine efficacy in hospitalized COVID-19 patients across various outcomes.** (a) Forest plot showing the risk difference (RD) for ICU admissions in patients receiving colchicine (n = 387) and those not receiving colchicine (n = 381). The pooled RD is -0.03 (95% CI: [-0.06, 0.01]), with no observed heterogeneity ($I^2$ = 0%). (b) Funnel plot assessing publication bias for studies reporting ICU admissions. (c) Forest plot showing the mean difference (MD) in hospitalization days between patients receiving colchicine (n = 8,208) and those not receiving colchicine (n = 8,321). The pooled MD is -0.58 days (95% CI: [-1.80, 0.64]), with high heterogeneity ($I^2$ = 68%). (d) Funnel plot assessing publication bias for studies reporting hospitalization days. (e) Forest plot showing the mean difference (MD) in ICU days between patients receiving colchicine (n = 8,208) and those not receiving colchicine (n = 8,321). The pooled MD is -1.41 days (95% CI: [-3.35, 0.53]), with moderate heterogeneity ($I^2$ = 32%). (f) Funnel plot assessing publication bias for studies reporting ICU days.

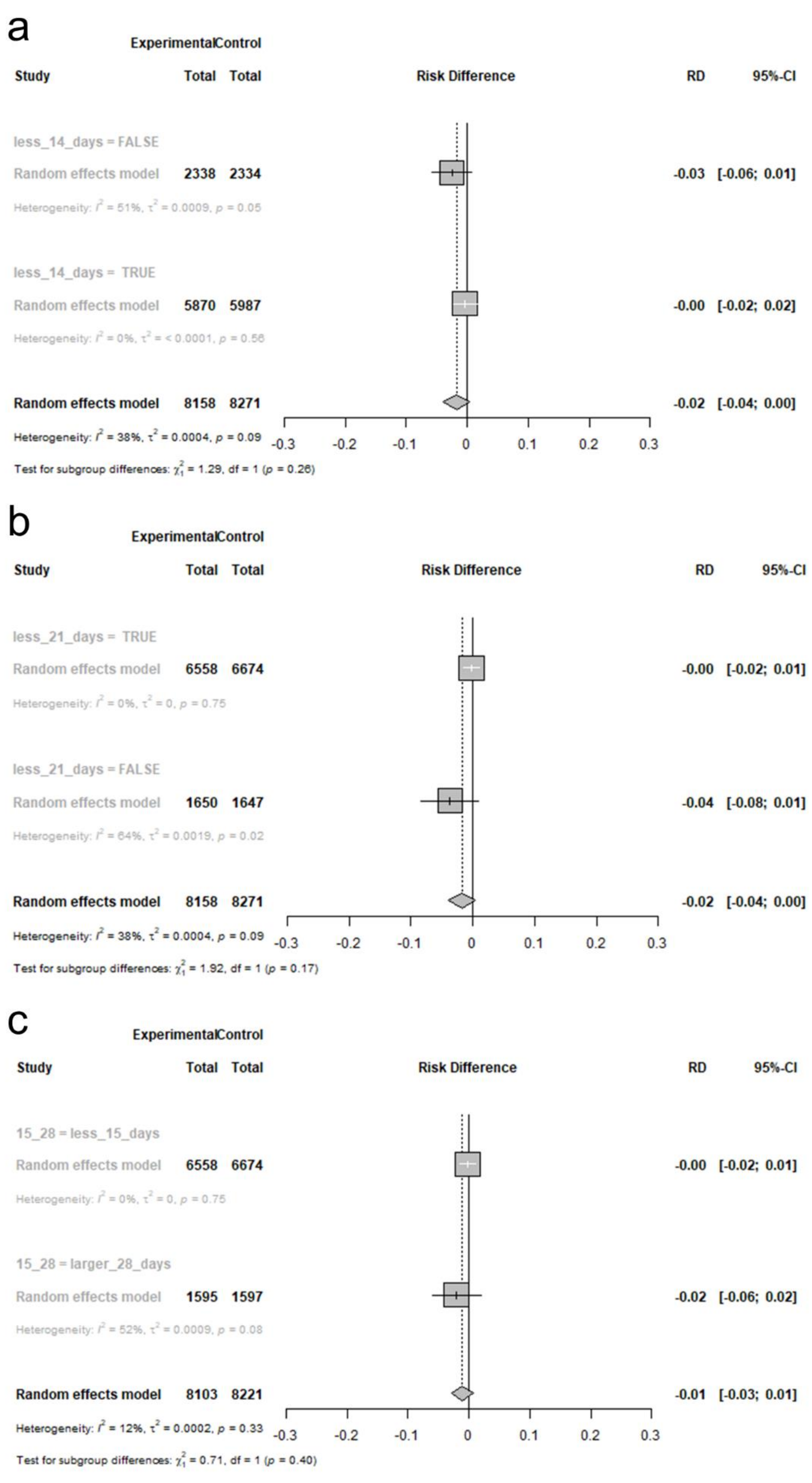
a
ExperimentalControl
Study Total Total Risk Difference RD 95%-CI
less_14_days = FALSE
Random effects model 2338 2334 -0.03 [-0.06; 0.01]
Heterogeneity: I² = 51%, τ² = 0.0009, p = 0.05
less_14_days = TRUE
Random effects model 5870 5987 -0.00 [-0.02; 0.02]
Heterogeneity: I² = 0%, τ² = < 0.0001, p = 0.56
Random effects model 8158 8271 -0.02 [-0.04; 0.00]
Heterogeneity: I² = 38%, τ² = 0.0004, p = 0.09
-0.3 -0.2 -0.1 0 0.1 0.2 0.3
Test for subgroup differences: χ²₁ = 1.29, df = 1 (p = 0.26)
b
ExperimentalControl
Study Total Total Risk Difference RD 95%-CI
less_21_days = TRUE
Random effects model 6558 6674 -0.00 [-0.02; 0.01]
Heterogeneity: I² = 0%, τ² = 0, p = 0.75
less_21_days = FALSE
Random effects model 1650 1647 -0.04 [-0.08; 0.01]
Heterogeneity: I² = 64%, τ² = 0.0019, p = 0.02
Random effects model 8158 8271 -0.02 [-0.04; 0.00]
Heterogeneity: I² = 38%, τ² = 0.0004, p = 0.09
-0.3 -0.2 -0.1 0 0.1 0.2 0.3
Test for subgroup differences: χ²₁ = 1.92, df = 1 (p = 0.17)
c
ExperimentalControl
Study Total Total Risk Difference RD 95%-CI
15_28 = less_15_days
Random effects model 6558 6674 -0.00 [-0.02; 0.01]
Heterogeneity: I² = 0%, τ² = 0, p = 0.75
15_28 = larger_28_days
Random effects model 1595 1597 -0.02 [-0.06; 0.02]
Heterogeneity: I² = 52%, τ² = 0.0009, p = 0.08
Random effects model 8103 8221 -0.01 [-0.03; 0.01]
Heterogeneity: I² = 12%, τ² = 0.0002, p = 0.33
-0.3 -0.2 -0.1 0 0.1 0.2 0.3
Test for subgroup differences: χ²₁ = 0.71, df = 1 (p = 0.40)

**Supplemental Figure 02. Subgroup meta-analysis of colchicine efficacy on mortality in hospitalized COVID-19 patients based on different time intervals.** (a) Forest plot showing the risk difference (RD) for mortality in patients receiving colchicine with different time frames: less than 14 days (n = 5,870) and more than 14 days (n = 2,338). The RD for mortality with <14 days was 0.00 (95% CI: [-0.02, 0.02], $I^2$ = 0%), and with >14 days was -0.03 (95% CI: [-0.06, 0.01], $I^2$ = 51%). (b) Forest plot of mortality risk difference in colchicine-treated patients within the first 21 days (n = 6,558) compared to those treated after 21 days (n = 1,650). The RD for mortality with <21 days was 0.00 (95% CI: [-0.02, 0.01], $I^2$ = 0%), while with >21 days was -0.04 (95% CI: [-0.08, 0.01], $I^2$ = 64%). (c) Forest plot of mortality risk difference in colchicine-treated patients within the first 15 days (n = 6,558) compared to those treated after 28 days (n = 1,595). The RD for mortality with <15 days was 0.00 (95% CI: [-0.02, 0.01], $I^2$ = 0%), and with >28 days was -0.02 (95% CI: [-0.06, 0.02], $I^2$ = 52%).

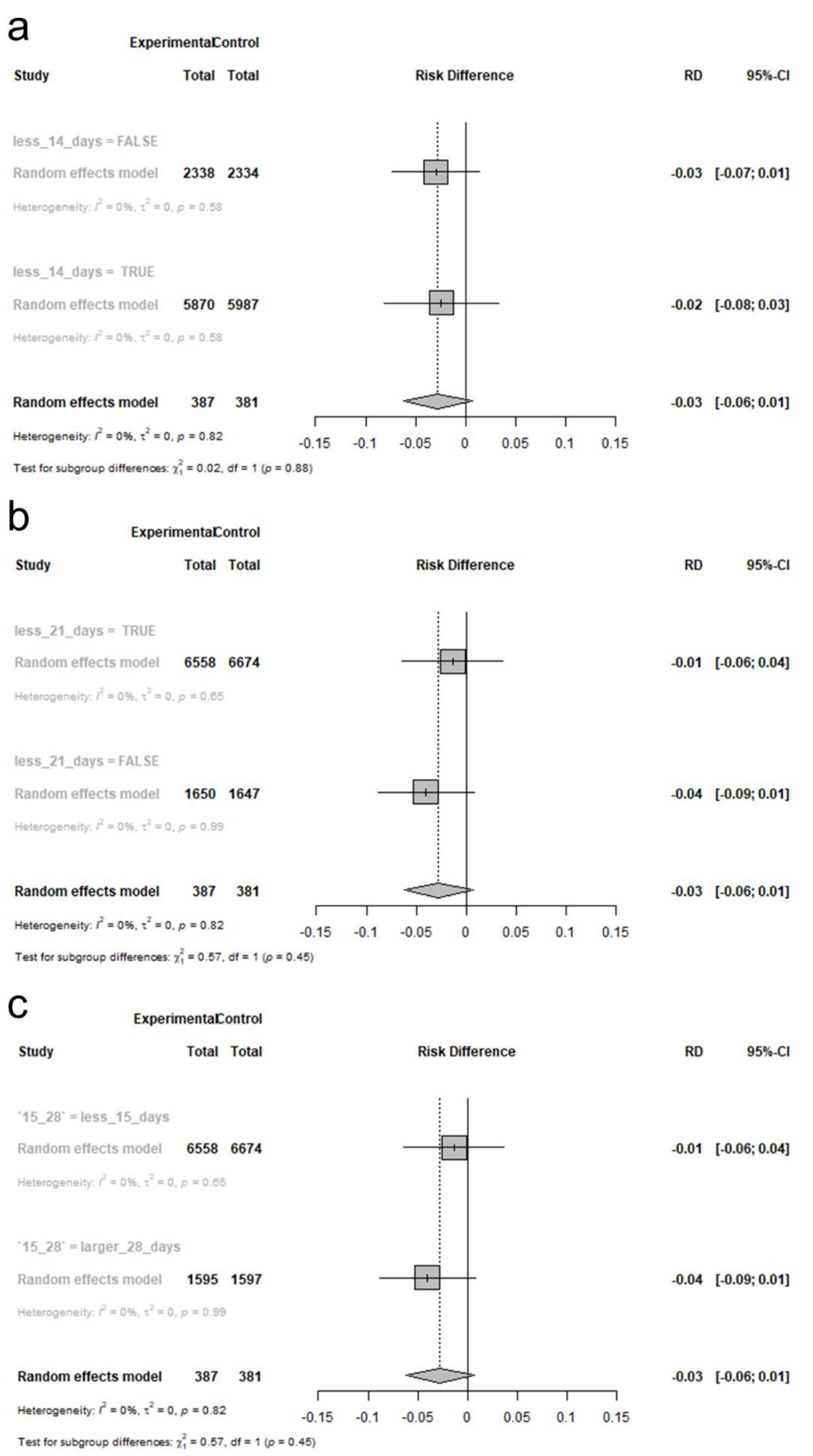
a
Experimental Control
Study Total Total Risk Difference RD 95%-CI
less_14_days = FALSE
Random effects model 2338 2334 -0.03 [-0.07; 0.01]
Heterogeneity: $I^2$ = 0%, $\tau^2$ = 0, $p$ = 0.58
less_14_days = TRUE
Random effects model 5870 5987 -0.02 [-0.08; 0.03]
Heterogeneity: $I^2$ = 0%, $\tau^2$ = 0, $p$ = 0.58
Random effects model 387 381 -0.03 [-0.06; 0.01]
Heterogeneity: $I^2$ = 0%, $\tau^2$ = 0, $p$ = 0.82
Test for subgroup differences: $\chi^2_1$ = 0.02, df = 1 ($p$ = 0.88)
-0.15 -0.1 -0.05 0 0.05 0.1 0.15
b
Experimental Control
Study Total Total Risk Difference RD 95%-CI
less_21_days = TRUE
Random effects model 6558 6674 -0.01 [-0.06; 0.04]
Heterogeneity: $I^2$ = 0%, $\tau^2$ = 0, $p$ = 0.65
less_21_days = FALSE
Random effects model 1650 1647 -0.04 [-0.09; 0.01]
Heterogeneity: $I^2$ = 0%, $\tau^2$ = 0, $p$ = 0.99
Random effects model 387 381 -0.03 [-0.06; 0.01]
Heterogeneity: $I^2$ = 0%, $\tau^2$ = 0, $p$ = 0.82
Test for subgroup differences: $\chi^2_1$ = 0.57, df = 1 ($p$ = 0.45)
-0.15 -0.1 -0.05 0 0.05 0.1 0.15
c
Experimental Control
Study Total Total Risk Difference RD 95%-CI
`15_28` = less_15_days
Random effects model 6558 6674 -0.01 [-0.06; 0.04]
Heterogeneity: $I^2$ = 0%, $\tau^2$ = 0, $p$ = 0.65
`15_28` = larger_28_days
Random effects model 1595 1597 -0.04 [-0.09; 0.01]
Heterogeneity: $I^2$ = 0%, $\tau^2$ = 0, $p$ = 0.99
Random effects model 387 381 -0.03 [-0.06; 0.01]
Heterogeneity: $I^2$ = 0%, $\tau^2$ = 0, $p$ = 0.82
Test for subgroup differences: $\chi^2_1$ = 0.57, df = 1 ($p$ = 0.45)
-0.15 -0.1 -0.05 0 0.05 0.1 0.15

**Supplemental Figure 03. Subgroup meta-analysis of colchicine efficacy on intensive care unit (ICU) admissions in hospitalized COVID-19 patients based on different time intervals.** (a) Forest plot showing the risk difference (RD) for ICU admissions in patients receiving colchicine with different time frames: less than 14 days (n = 5,870) and more than 14 days (n = 2,338). The RD for ICU admissions with <14 days was -0.02 (95% CI: [-0.08, 0.03], $I^2$ = 0%), and with >14 days was -0.03 (95% CI: [-0.07, 0.01], $I^2$ = 0%). (b) Forest plot of ICU admissions risk difference in colchicine-treated patients within the first 21 days (n = 6,558) compared to those treated after 21 days (n = 1,650). The RD for ICU admissions with <21 days was -0.01 (95% CI: [-0.06, 0.04], $I^2$ = 0%), and with >21 days was -0.04 (95% CI: [-0.09, 0.01], $I^2$ = 0%). (c) Forest plot of ICU admissions risk difference in colchicine-treated patients within the first 15 days (n = 6,558) compared to those treated after 28 days (n = 1,595). The RD for ICU admissions with <15 days was -0.01 (95% CI: [-0.06, 0.04], $I^2$ = 0%), and with >28 days was -0.04 (95% CI: [-0.09, 0.01], $I^2$ = 0%).

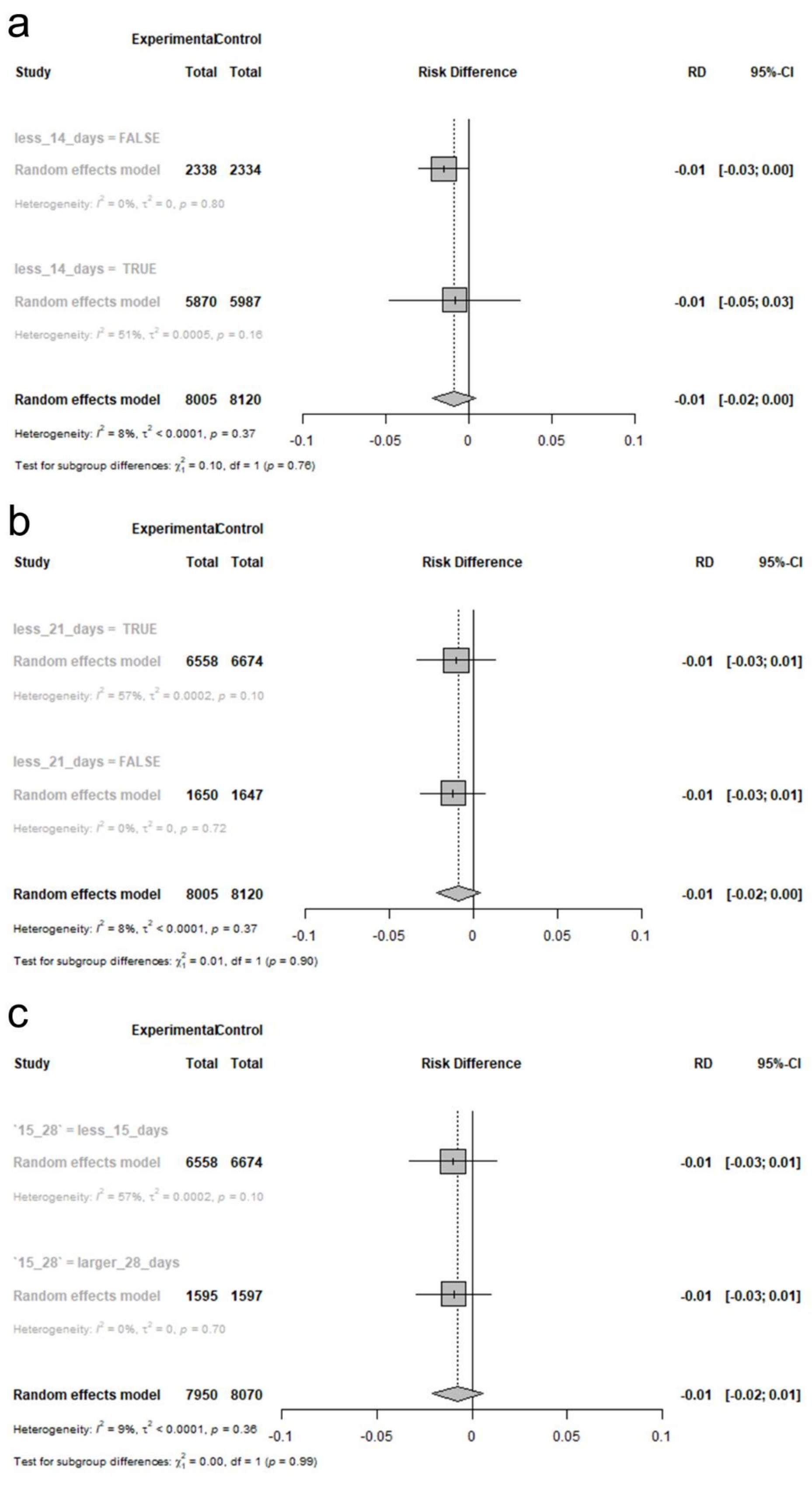

a
Experimental Control
Study Total Total Risk Difference RD 95%-CI
less_14_days = FALSE
Random effects model 2338 2334 -0.01 [-0.03; 0.00]
Heterogeneity: $I^2$ = 0%, $\tau^2$ = 0, $p$ = 0.80
less_14_days = TRUE
Random effects model 5870 5987 -0.01 [-0.05; 0.03]
Heterogeneity: $I^2$ = 51%, $\tau^2$ = 0.0005, $p$ = 0.16
Random effects model 8005 8120 -0.01 [-0.02; 0.00]
Heterogeneity: $I^2$ = 8%, $\tau^2$ < 0.0001, $p$ = 0.37
Test for subgroup differences: $\chi^2_1$ = 0.10, df = 1 ($p$ = 0.76)
-0.1 -0.05 0 0.05 0.1
b
Experimental Control
Study Total Total Risk Difference RD 95%-CI
less_21_days = TRUE
Random effects model 6558 6674 -0.01 [-0.03; 0.01]
Heterogeneity: $I^2$ = 57%, $\tau^2$ = 0.0002, $p$ = 0.10
less_21_days = FALSE
Random effects model 1650 1647 -0.01 [-0.03; 0.01]
Heterogeneity: $I^2$ = 0%, $\tau^2$ = 0, $p$ = 0.72
Random effects model 8005 8120 -0.01 [-0.02; 0.00]
Heterogeneity: $I^2$ = 8%, $\tau^2$ < 0.0001, $p$ = 0.37
Test for subgroup differences: $\chi^2_1$ = 0.01, df = 1 ($p$ = 0.90)
-0.1 -0.05 0 0.05 0.1
c
Experimental Control
Study Total Total Risk Difference RD 95%-CI
`15_28` = less_15_days
Random effects model 6558 6674 -0.01 [-0.03; 0.01]
Heterogeneity: $I^2$ = 57%, $\tau^2$ = 0.0002, $p$ = 0.10
`15_28` = larger_28_days
Random effects model 1595 1597 -0.01 [-0.03; 0.01]
Heterogeneity: $I^2$ = 0%, $\tau^2$ = 0, $p$ = 0.70
Random effects model 7950 8070 -0.01 [-0.02; 0.01]
Heterogeneity: $I^2$ = 9%, $\tau^2$ < 0.0001, $p$ = 0.36
Test for subgroup differences: $\chi^2_1$ = 0.00, df = 1 ($p$ = 0.99)
-0.1 -0.05 0 0.05 0.1

**Supplemental Figure 04. Subgroup meta-analysis of colchicine efficacy on mechanical ventilation (MV) in hospitalized COVID-19 patients based on different time intervals.** (a) Forest plot showing the risk difference (RD) for mechanical ventilation in patients receiving colchicine with different time frames: less than 14 days (n = 5,870) and more than 14 days (n = 2,338). The RD for MV with <14 days was -0.01 (95% CI: [-0.05, 0.03], $I^2$ = 51%), and with >14 days was -0.01 (95% CI: [-0.03, 0.00], $I^2$ = 0%). (b) Forest plot of mechanical ventilation risk difference in colchicine-treated patients within the first 21 days (n = 6,558) compared to those treated after 21 days (n = 1,650). The RD for MV with <21 days was -0.01 (95% CI: [-0.03, 0.01], $I^2$ = 57%), and with >21 days was -0.01 (95% CI: [-0.03, 0.01], $I^2$ = 0%). (c) Forest plot of mechanical ventilation risk difference in colchicine-treated patients within the first 15 days (n = 6,558) compared to those treated after 28 days (n = 1,595). The RD for MV with <15 days was -0.01 (95% CI: [-0.03, 0.01], $I^2$ = 57%), and with >28 days was -0.01 (95% CI: [-0.03, 0.01], $I^2$ = 0%).

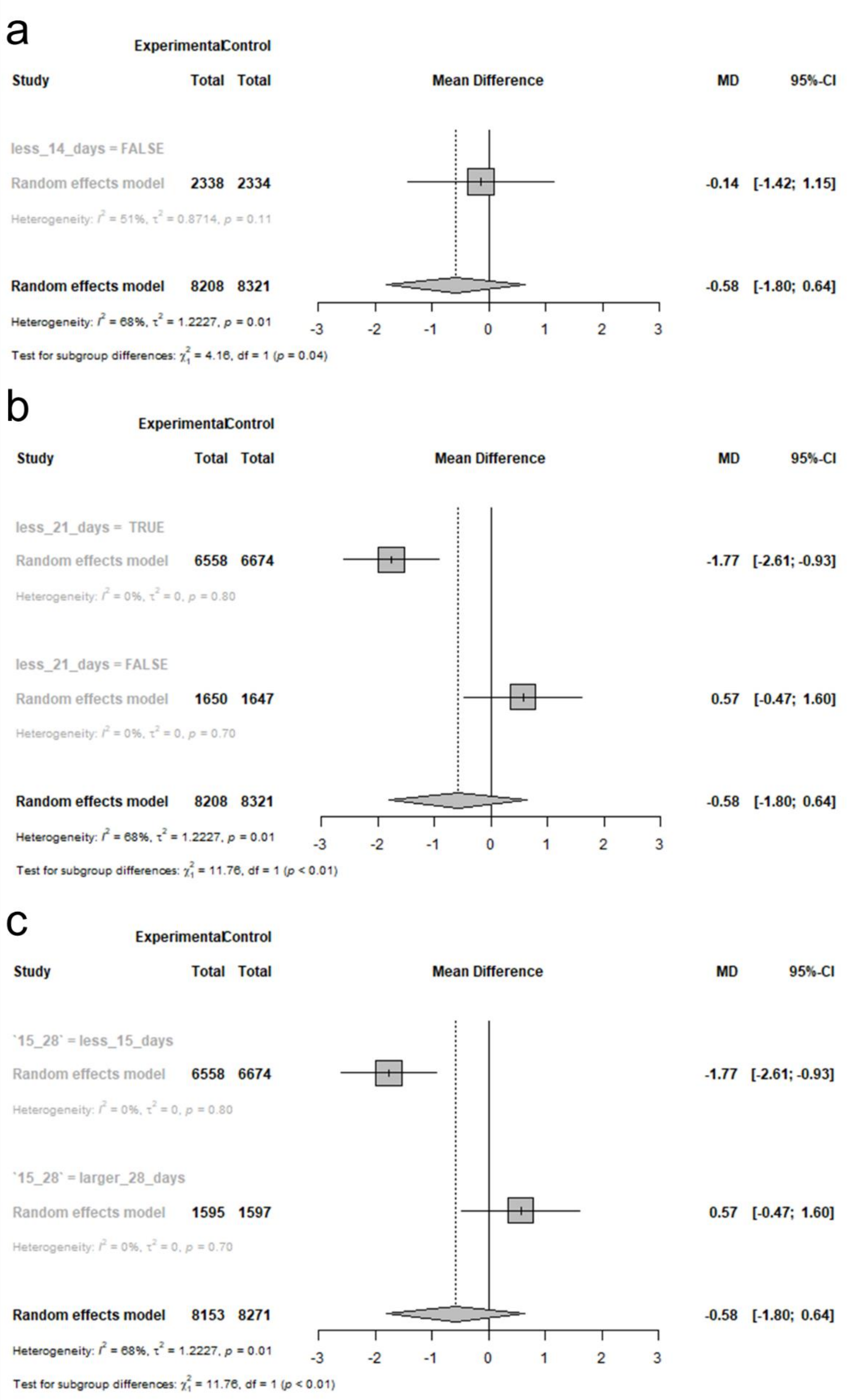
a
Experimental Control
Study
Total Total
Mean Difference
MD
95%-CI
less_14_days = FALSE
Random effects model 2338 2334
-0.14 [-1.42; 1.15]
Heterogeneity: I^2 = 51%, τ^2 = 0.8714, p = 0.11
Random effects model 8208 8321
-0.58 [-1.80; 0.64]
Heterogeneity: I^2 = 68%, τ^2 = 1.2227, p = 0.01
Test for subgroup differences: χ^2_1 = 4.16, df = 1 (p = 0.04)
-3 -2 -1 0 1 2 3
b
Experimental Control
Study
Total Total
Mean Difference
MD
95%-CI
less_21_days = TRUE
Random effects model 6558 6674
-1.77 [-2.61; -0.93]
Heterogeneity: I^2 = 0%, τ^2 = 0, p = 0.80
less_21_days = FALSE
Random effects model 1650 1647
0.57 [-0.47; 1.60]
Heterogeneity: I^2 = 0%, τ^2 = 0, p = 0.70
Random effects model 8208 8321
-0.58 [-1.80; 0.64]
Heterogeneity: I^2 = 68%, τ^2 = 1.2227, p = 0.01
Test for subgroup differences: χ^2_1 = 11.76, df = 1 (p < 0.01)
-3 -2 -1 0 1 2 3
c
Experimental Control
Study
Total Total
Mean Difference
MD
95%-CI
`15_28` = less_15_days
Random effects model 6558 6674
-1.77 [-2.61; -0.93]
Heterogeneity: I^2 = 0%, τ^2 = 0, p = 0.80
`15_28` = larger_28_days
Random effects model 1595 1597
0.57 [-0.47; 1.60]
Heterogeneity: I^2 = 0%, τ^2 = 0, p = 0.70
Random effects model 8153 8271
-0.58 [-1.80; 0.64]
Heterogeneity: I^2 = 68%, τ^2 = 1.2227, p = 0.01
Test for subgroup differences: χ^2_1 = 11.76, df = 1 (p < 0.01)
-3 -2 -1 0 1 2 3

**Supplemental Figure 05. Subgroup meta-analysis of colchicine efficacy on hospitalization in hospitalized COVID-19 patients based on different time intervals.** (a) Forest plot showing the mean difference (MD) for hospitalization in patients receiving colchicine with different time frames: more than 14 days (n = 2,338). The MD for hospitalization with >14 days was -0.14 (95% CI: [-1.42, 1.15], $I^2$ = 51%). (b) Forest plot of hospitalization mean difference in colchicine-treated patients within the first 21 days (n = 6,558) compared to those treated after 21 days (n = 1,650). The MD for hospitalization with <21 days was -1.77 (95% CI: [-2.61, -0.93], $I^2$ = 0%), and with >21 days was 0.57 (95% CI: [-0.47, 1.60], $I^2$ = 0%). (c) Forest plot of hospitalization mean difference in colchicine-treated patients within the first 15 days (n = 6,558) compared to those treated after 28 days (n = 1,595). The MD for hospitalization with <15 days was -1.77 (95% CI: [-2.61, -0.93], $I^2$ = 0%), and with >28 days was 0.57 (95% CI: [-0.47, 1.60], $I^2$ = 0%).

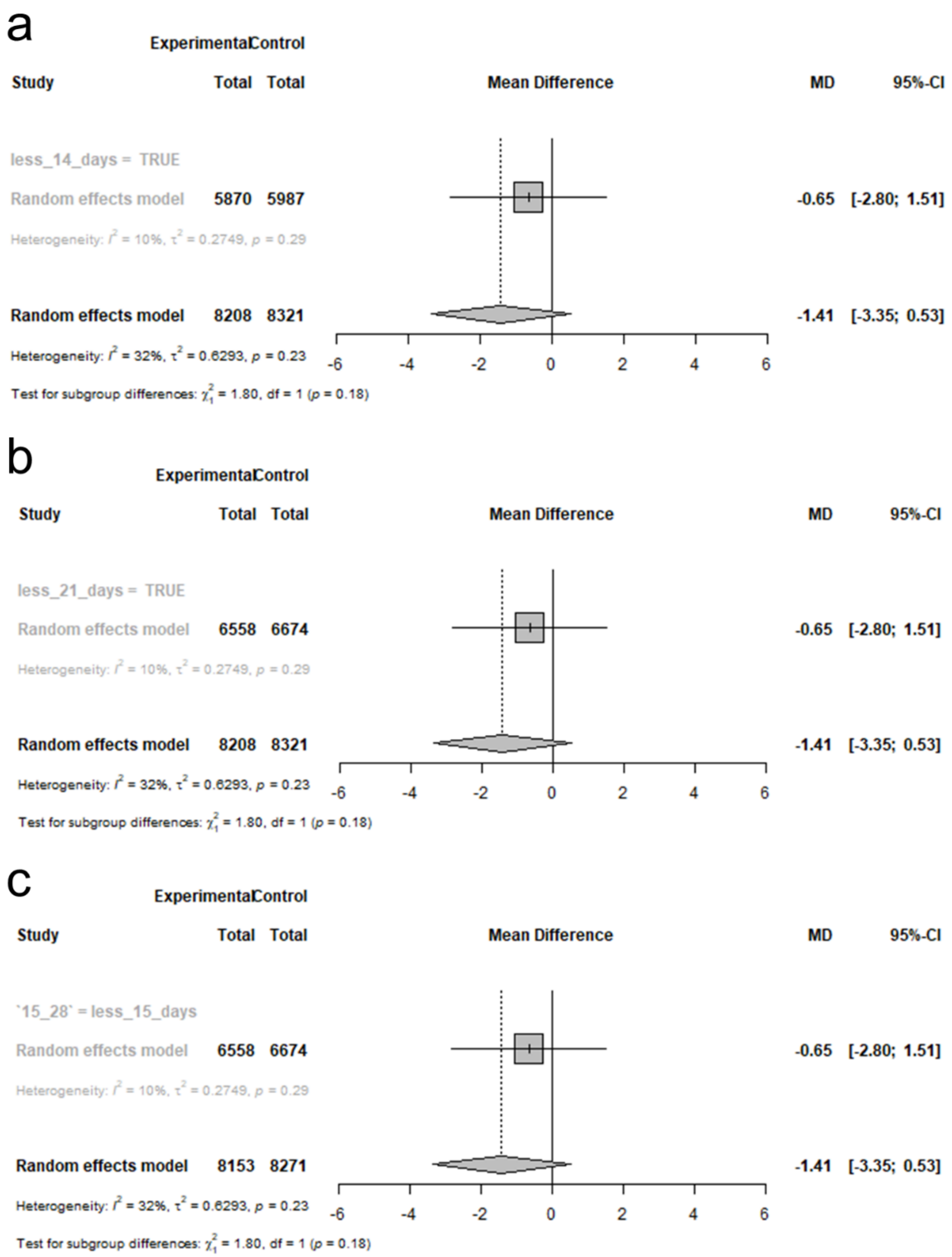


**Supplemental Figure 06. Subgroup meta-analysis of colchicine efficacy on ICU days in hospitalized COVID-19 patients based on different time intervals.** (a) Forest plot showing the mean difference (MD) in ICU days for patients receiving colchicine with different time frames: less than 14 days (n = 5,870). The MD for ICU days with <14 days was -0.65 (95% CI:

[-2.80, 1.51], $I^2 = 10\%$). (b) Forest plot of ICU days mean difference in colchicine-treated patients within the first 21 days (n = 6,558) compared to those treated after 21 days (n = 1,650). The MD for ICU days with <21 days was -0.65 (95% CI: [-2.80, 1.51], $I^2 = 10\%$). (c) Forest plot of ICU days mean difference in colchicine-treated patients within the first 15 days (n = 6,558) compared to those treated after 28 days (n = 1,595). The MD for ICU days with <15 days was -0.65 (95% CI: [-2.80, 1.51], $I^2 = 10\%$).
